# Why Women Choose At-Home Abortion via Teleconsultation in France?: A Mixed-Method Study on Drivers of Telemedicine Abortion

**DOI:** 10.1101/2021.04.19.21255757

**Authors:** Hazal Atay, Helene Perivier, Kristina Gemzell-Danielson, Jean Guilleminot, Danielle Hassoun, Judith Hottois, Rebecca Gomperts, Emmanuelle Levrier

**Author notes:** **Corresponding author** Hazal ATAY, CEVIPOF, Sciences Po Paris, 98 rue de l’Université 75007 Paris.

## Abstract

**Objectives:** In face of the COVID-19 health emergency, France has allowed medical abortions to be performed by teleconsultation until 9 weeks of gestation. In an attempt to understand the demand and main drivers of telemedicine abortion, we analysed the requests that Women on Web (WoW), an online telemedicine abortion service operating worldwide, received from France throughout 2020.

**Methods:** We conducted a parallel convergent mixed-method study among 809 consultations received from France at WoW between 1 January and 31 December 2020. We performed a cross-sectional study of data obtained from the WoW consultation survey and a manifest content analysis of anonymised email correspondence of 140 women consulting with WoW helpdesk from France.

**Results:** We found that women encounter macro-level, individual-level, and provider-level constraints while trying to access abortion in France. The preferences and needs over secrecy (46.2%), privacy (38.3%), and comfort (34.9%) are among the most frequent reasons for women from France to choose telemedicine abortion through WoW. The COVID-19 pandemic seems to be an important driver for resorting to telemedicine (30.6%). The lockdowns seem to have had an impact on the number of consultations received at WoW from France, increasing from 60 in March to 128 in April during the first lockdown and from 54 in October to 80 in November during the second lockdown.

**Conclusions:** The demand for at-home medical abortion via teleconsultation increased in France during the lockdowns. However, drivers of telemedicine abortion are multi-dimensional and go beyond the conditions unique to the pandemic. Given the various constraints women continue to encounter in accessing safe abortion, telemedicine can help meet women’s preferences and needs for secrecy, privacy and comfort, while facilitating improved access to and enabling more person-centred abortion care.

**Tweetable Abstract:** At-home abortion via teleconsultation can help meet women’s needs and preferences for privacy, secrecy, and comfort, while facilitating improved access to abortion care in France.

**Key Messages:**

- The lockdowns seem to have had an impact on the number of consultations received at WoW from France, increasing from 60 in March to 128 in April during the first lockdown and from 54 in October to 80 in November during the second lockdown.
- While the COVID-19 pandemic was an important push factor for women to choose telemedicine, the drivers of telemedicine are multidimensional and go beyond conditions unique to the pandemic.
- Telemedicine can help meet women’s needs and preferences for privacy, secrecy, and comfort, while facilitating access to and enabling more person-centred abortion care in France.

**Funding Information:** This research was funded by a public grant overseen by the French National Research Agency (ANR) as part of the “Investissements d’Avenir” program LIEPP (ANR-11-LABX-0091, ANR-11-IDEX-0005-02) and the Université de Paris IdEx (ANR-18-IDEX-0001).

**Patient and public involvement statement:** This public policy analysis does not involve patients or the public in the design, or conduct, or reporting, or dissemination plans of this work. However, the service that WoW provides is designed to address the priorities and experiences of people who access the service. Thus, the research questions were informed by the needs of people who rely on WoW to access abortion.

**Ethics approval:** The study was approved by the Regional Ethics Committee, Karolinska Institutet, Dnr 2009/2072-31/2 and Dnr 2020/05406.

## Introduction

The COVID-19 outbreak has posed significant challenges for the provision of abortion care. In the context of the pandemic, national abortion guidelines in France have changed several times. In April 2020, France introduced a temporary exemption from the requirement to take the abortion medication in presence of a medical doctor or midwife and allowed at-home medical abortion via teleconsultation until 7 weeks of pregnancy via teleconsultation [1]. The measure was adopted in the midst of a national lockdown, which started on 17 March 2020 and continued until 11 May 2020, and was justified as per the recommendations of the World Health Organization (WHO), advances in telemedicine, and the “strong mobilization of health establishment in the management of the crisis and the need to limit consultations in hospitals for any other reason” [1]. The measure allowed women to book appointments online, purchase both abortion pills, mifepristone and misoprostol, from a pharmacy on prescription, and self-manage their abortions at home. Abandoned in July, the measure was once again embraced in November as the health crisis intensified, this time extending the gestational age for at-home medical abortion via teleconsultation from 7 weeks to 9 weeks upon the recommendations of the High Authority of Health on “Rapid Responses COVID-19: Voluntary Interruption of Pregnancy (IVG)” [2]. It was affirmed that medical abortion pills can be delivered directly to individuals in pharmacies with an “exceptional delivery” (*délivrance exceptionnelle*) notation. Similar to the previous measure, the decision was adopted in the middle of a (second) nation-wide lockdown, which started on 30 October and continued until 15 December 2020.

The French abortion guidelines allow abortions to be performed until 14 weeks of gestation. Since 2013, abortion in public hospitals is fully reimbursed by the French social security for all women, including undocumented immigrants [3]. Since 2016, midwives are authorized to practice medical abortions and every procedure required within the framework of abortion care, including ultrasound and post-abortion care, is fully reimbursed by the social security [4]. However, prior to the COVID-19 pandemic, telemedicine abortion provision was not allowed; individuals were instead required to take the first abortion medication, mifepristone, in presence of a medical doctor or a midwife [5]. Medical abortions until 7 weeks of gestation were performed in private offices of doctors and midwives, primary care centres (*centre de santé*), family planning centres or at the hospitals. Between 7 and 9 weeks of gestation, medical abortion was performed only in hospital settings; women were required to take the first abortion medication, mifepristone, at the hospital and had to come back 48 hours later to take the second medication, misoprostol. They were then required to stay at the hospital for 3 hours following the intake of the misoprostol [6].

France was not the only country in Europe to reconsider the regulations governing the dispensation of medical abortion pills during the pandemic. Similar measures were also adopted in England, Wales, Scotland, and Ireland [7]. These measures were often based on the WHO guidelines, recommending telemedicine and self-care interventions for the provision of medical abortion within the first trimester (12 weeks of gestation) [8]. The WHO contends that at-home abortion via teleconsultation is acceptable, non-invasive and cost effective [8]. It further suggests that self-management of abortion at home improves autonomy, by enabling a sense of control over one’s own body and the abortion procedure [8]. In fact, rights groups and experts have previously challenged the “overregulation” of the abortion pills [9] and have been calling for abortion pills to be available in pharmacies [10] and for home medical abortion to be provided via telemedicine prior to the pandemic [11]. Given that certain countries facilitated expanded access to abortion pills with the COVID-19 outbreak, several groups and practitioners now call for the adopted measures to continue beyond the pandemic [12, 13].

Endorsed by the WHO, telemedicine came to the fore as the silver lining amid the COVID-19 pandemic [14]. In light of these recent changes and debates, we examined why women chose at-home abortion via teleconsultation in France. With this objective, we analysed help requests that Women on Web (WoW), an online telemedicine abortion service operating worldwide, received from France throughout 2020.

## Methods

We conducted a parallel convergent mixed-method study among the 809 consultations received from France at WoW between 1 January and 31 December 2020. Within the framework of this research, we analysed two main data components. The first component is the data obtained from WoW consultation questionnaire and the second is the data obtained from women’s anonymised email correspondence with the WoW helpdesk. Both components were collected simultaneously and analysed independently. They were then merged to validate data and results and to interpret the findings better.

### Cross-Sectional Analysis of the Survey Data

We performed a cross-sectional study of the survey data obtained from WoW consultation survey that participants completed while requesting help online. The survey consists of 25 questions and is available online on WoW homepage [15]. The WoW consultation process has been described previously [16]. As part of the survey, participants gave their informed consent for their data to be used anonymously for research purposes [17]. The survey consists of both categorical and continuous data; continuous data was summarised in median and interquartile range, and categorical data in frequencies. We distinguished categorical data as per age groups to map demographic patterns. Moreover, we also distinguished between consultations where COVID-19 was mentioned and not mentioned among reasons, to suggest a counterfactual effect for reasons within and outside the pandemic context. The data analysis was done with IBM SPSS Statistics for Macintosh, Version 27.0 (IBM Corp. Released 2020).

### Content Analysis of Email Correspondence

When women from France completed their online WoW consultations, they were given information on local abortion services and were asked:

> If you are unable to access abortion services in France, could you please tell us a bit more about why? […] We will let you know as soon as possible if we can help you in any way.

In order to better understand women’s motivations for choosing telemedicine, and also to map the perceived barriers of access to abortion in France, we conducted a manifest content analysis of women’s email correspondence. Not all women who filled in the online consultation at WoW website proceeded with email correspondence. Two researchers (HA and EL) analysed the email correspondence of those who did until data saturation, when new data started to be redundant of data already collected [18]. In total, 157 emails were analysed at manifest level from correspondence with 140 women. Emails varied in length from 4 to 540 words. The content analysis was conducted in QSR international Pty Ltd. NVivo (released in March 2020). All researchers contributed to the identification and categorisation of the recurring themes. The categories were then quantified and merged with the findings of the cross-sectional analysis for interpretation.

## Results

### Cross-Sectional Study

Women on Web opened its service for France on 1 January 2020 for the purpose of this research. From 1 January to 31 December 2020, 809 women from France completed online consultation on the WoW website in which they requested telemedicine abortion services. This time period has also coincided with a highly challenging health emergency: the COVID-19 pandemic. Throughout 2020, the French government took several measures to reduce the spread and transmissions of the coronavirus, including nation-wide lockdowns: first from 17 March to 11 May and second from 30 October to 15 December 2020. Lockdowns entailed closure of borders and strict travel restrictions. It is in this context that the Figure 1 represents the number of telemedicine abortion consultations received from France at WoW during 2020.

**Figure 1:**
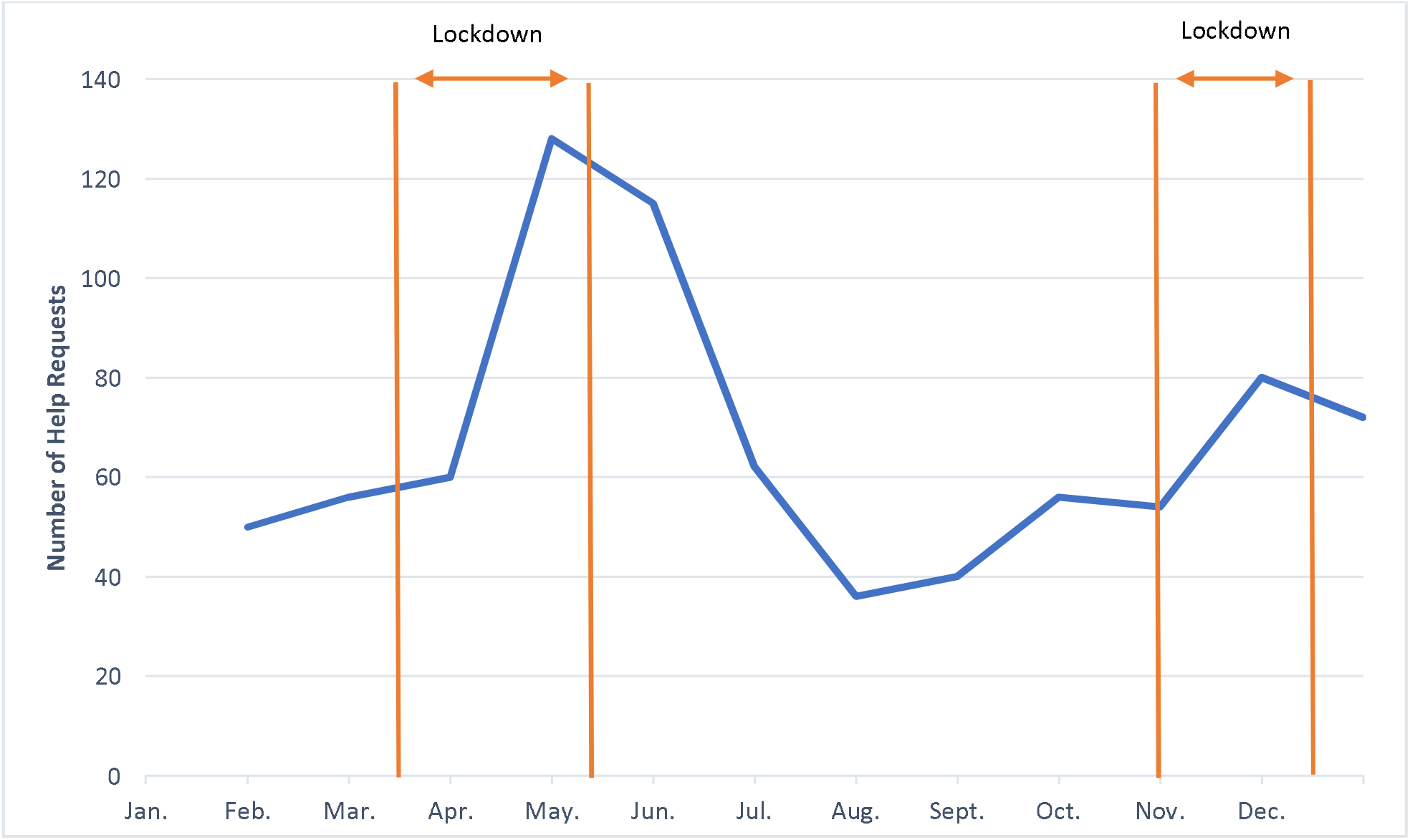
The Number of Telemedical Abortion Consultations from France received by Women on Web between 1 January and 31 December 2020 (n = 809).

**Figure 2:**
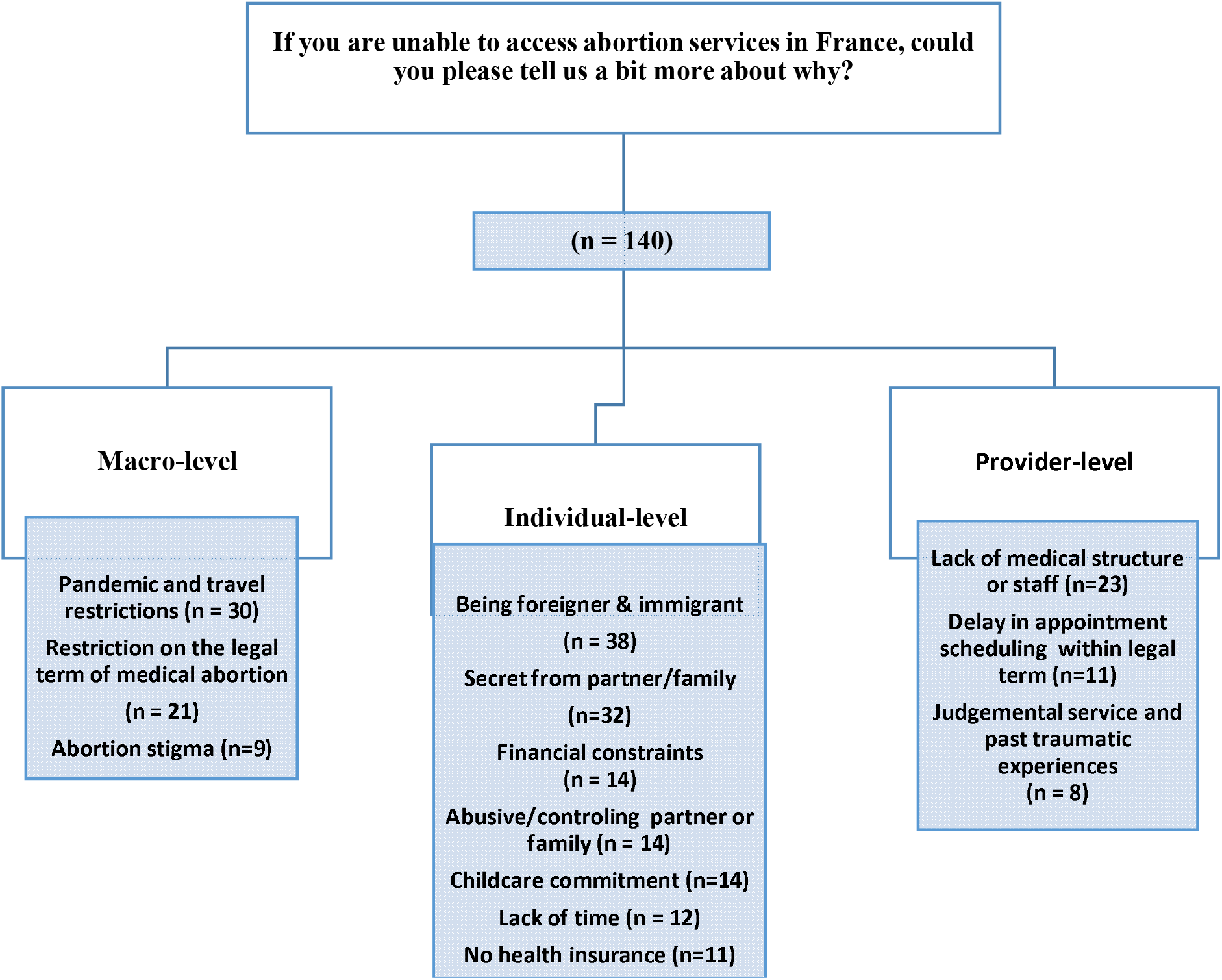
Schematic presentation of categories and subcategories derived from email correspondence of 140 women from France who consulted Women on Web between 1 January and 31 December 2020. *Women often had multiple constraints that cut across different categories and subcategories, and therefore the sum of correspondents for all categories exceeds the overall number of respondents.

Table 1 summarises background and pregnancy & abortion related experiences of women. Table 2 illustrates the reasons why women from France requested telemedicine abortion through WoW.

**Table 1:** Background and pregnancy and abortion-related experiences of women from France requesting telemedicine abortion through WoW between 1 January and 31 December 2020.

| Demographic data | Median (IQR) |
| --- | --- |
| Age (in years) | 29 (11) |
| Gestational Age (in days) | 36 (15) |
| **Ultrasound prior to consultation (n = 774, missing n = 35)** | **Frequency in % (n)** |
| Yes | 16.7 (130) |
| No | 83.2 (644) |
| **Reasons for not having an ultrasound (n = 661, missing n = 148)** |  |
| I am afraid my partner or other people will find out | 32.9 (218) |
| I thought I did not need one as I am sure I am pregnant, and I know how long I have been pregnant | 32.0 (212) |
| I cannot afford one | 31.6 (209) |
| I cannot get to a clinic to get one because of distance or lack of transportation | 16.4 (109) |
| I just did not have time to do it | 13.0 (86) |
| I did not know that I needed one | 7.4 (49) |
| I am unsure where to get one | 7.4 (49) |
| **Reasons for unwanted pregnancy (n = 809)** |  |
| Contraceptives that I used did not work | 53.8 (436) |
| Did not use any contraceptives | 41.4 (335) |
| Rape | 4.4 (36) |
| I wanted a pregnancy, but my situation has changed | 0.2 (2) |
| **Reasons for wanting an abortion (n = 766, missing n = 43)** |  |
| I just cannot raise a child at this point of my life | 53.1 (407) |
| No money to raise a child | 41.8 (328) |
| My family is complete | 27.4 (210) |
| I am too young | 15.4 (118) |
| Want to finish school | 15.2 (117) |
| I am too old | 6.3 (49) |
| My partner does not want a child | 2.0 (16) |
| I am ill | 1.5 (12) |
| **Previous abortion experience (n = 737, missing n = 72)** |  |
| None | 64.4 (475) |
| 1 | 27.2 (201) |
| 2 or more | 8.2 (61) |
| **Child status (n = 750, missing n = 59)** |  |
| None | 42.1 (316) |
| 1 | 18.0 (135) |
| 2-3 children | 34.1 (256) |
| 4 or more | 5.7 (43) |

\* Multiple responses are allowed for the questions on reasons for not having an ultrasound and for wanting an abortion; therefore, total response exceeds 100%.
\* Questions regarding reasons for wanting an abortion, previous abortion experience, and child status are optional, which explains the missing data.

**Table 2:** Reasons Why Women Chose Telemedicine through WoW between 1 January and 31 December 2020 per Age Groups and COVID-19 Relevancy

| Frequencies in % (n) |  |  |  |
| --- | --- | --- | --- |
|  | Age groups | COVID-19 Relevancy | Total |

| **Reasons why women choose telemedicine abortion through Women on Web** | **18-25**<br>**(n = 261,**<br>**reasons missing**<br>**n=16)** | **26-35**<br>**(n = 344,**<br>**reasons missing**<br>**n=14)** | **36 and above**<br>**(n = 164,**<br>**reasons missing**<br>**n=10)** | **COVID-19**<br>**Related**<br>**Consultations**<br>**(n=236)** | **COVID-19**<br>**Unrelated**<br>**Consultations**<br>**(n=573)** | **(n=769, reasons**<br>**missing n=40)** |
| --- | --- | --- | --- | --- | --- | --- |
| Need to keep abortion a secret from partner or family | 58.2 (152) | 38.5 (138) | 37.9 (66) | 31.3 (74) | 49.2 (282) | 46.2 (356) |
| I would rather keep my abortion private | 44.8 (117) | 32.1 (115) | 36.2 (63) | 32.2 (76) | 38.2 (219) | 38.3 (295) |
| I would be more comfortable at home | 39.8 (104) | 28.7 (103) | 35.6 (62) | 34.7 (82) | 32.6 (187) | 34.9 (269) |
| Coronavirus | 35.6 (93) | 26.8 (96) | 37 (47) | 100 (236) | 0 (0) | 30.6 (236) |
| I would rather take care of my own abortion | 34.4 (90) | 26.2 (94) | 20.6 (36) | 0 (0) | 38.3 (220) | 28.6 (220) |
| It's hard to access abortion due to work or school commitments | 29.5 (77) | 20 (69) | 21.3 (35) | 16.52 (39) | 24.7 (142) | 23.5 (181) |
| It is hard to access abortion because of the cost | 31.4 (82) | 14.5 (52) | 12.6 (22) | 25.0 (39) | 20.4 (117) | 20.2 (156) |
| It is hard to access abortion because of childcare | 11.4 (30) | 21.8 (75) | 25.6 (42) | 16.10 (38) | 19.0 (109) | 19.1 (147) |
| I would rather have my partner or friend with me during the process | 25.2 (66) | 9.2 (33) | 8.6 (15) | 13.1 (31) | 14.4 (83) | 14.8 (114) |
| It is hard to access abortion because of legal restrictions | 11.1 (29) | 11.7 (42) | 8 (14) | 11.8 (28) | 9.9 (57) | 11 (85) |
| Stigma | 16 (42) | 8.3 (30) | 7.4 (13) | 11.0 (26) | 10.2 (59) | 11 (85) |
| It is hard to access abortion because of distance | 12.6 (33) | 5.2 (18) | 10.9 (18) | 11.0 (26) | 7.5 (43) | 8.9 (69) |
| Other reason | 6.1 (16) | 3.7 (13) | 7.3 (12) | 5.9 (14) | 4.7 (27) | 5.3 (41) |
| Abusive partner | 5.3 (14) | 4 (14) | 4.2 (7) | 3.8 (9) | 4.5 (26) | 4.5 (35) |
| I find it empowering | 4.9 (13) | 3.6 (13) | 2.2 (4) | 3.8 (9) | 3.6 (21) | 3.9 (30) |
| Undocumented immigrant | 3.4 (9) | 3.1 (11) | 3.6 (6) | 1.6 (4) | 3.8 (22) | 3.3 (26) |

\* The exact question reads “What are the main reasons why you are requesting an abortion through Women on Web?”
\* The question is optional, which explains the missing data.
\* Multiple responses are allowed; total response therefore exceeds 100%.

### Content Analysis

We grouped women’s replies in three main categories: macro-level constraints, individual-level constraints, and provider-level constraints. Macro-level constraints include socio-political conditions and legal frameworks; individual-level constraints concern women’s personal circumstances and preferences; and finally, provider-level constraints entail issues raised around service provision and access to available care.

### Integrated Results

The COVID-19 pandemic and associated lockdown measures restricted mobility, thereby adversely affecting availability of services. The lockdowns seem to have had an impact on th number of consultations received at WoW from France, increasing from 60 in March to 128 in April during the first lockdown and from 54 in October to 80 in November during the second lockdown. In their email correspondence, women reported not being able to access abortion care in France due to the pandemic and travel restrictions. As existing health structures were overburdened by the ongoing health emergency, women wrote about not being able to find or travel to an available medical structure or practitioner for abortion. A young woman’s email epitomises this situation:

> I can’t get an abortion as my family doesn’t know that I’m pregnant, and if they do find out, I’m in big trouble. I live in a village and it isn’t actually not possible for me to access abortion unless I travel to Switzerland, which is not an option now due to lockdown. The closest clinic is 2 hours away and I cannot go that far.

Delays in scheduling appointments, combined with the restriction on the legal term for medical abortion, appeared to be a frequent concern among many women consulting WoW. Several emails involve women contacting local clinics and associations and not being able to schedule an appointment within the legal gestational limit for medical abortion. A woman who was in such a situation wrote, “I want to remain in the legal delay for medical abortion, but I could not find any place to go before two months. I will pass the delay by then!”

While the lockdown measures increased the demand for telemedicine abortion, the drivers of telemedicine are in fact multi-dimensional and go beyond the unique conditions related to the pandemic. The preferences and needs over secrecy (46.2%), privacy (38.3 %), and comfort (34.9%), followed by coronavirus pandemic (30.6%), were among the most frequent reasons for women to choose telemedicine abortion in France. Women also indicated preference to take care of their own abortions as their reasons of choosing telemedicine through WoW, demonstrating a salient willingness to self-manage their abortion (28.6%). We found that similar frequencies, with slight fluctuations, are observed among COVID-19 related and unrelated consultations with the exception of willingness for self-management which appears to be exclusive to COVID-19 unrelated consultations. We observed that most of the time, women experience multiple constraints at the same time, which later informs their preferences; a woman requesting service wrote:

> I’m writing to you after being refused by 3 gynaecologists, they all refer me to the hospital which is overburdened with COVID. I contacted the family planning clinic, one is closed today, the other does not want to take me because I am not from their city. Besides the family planning clinic is difficult, they ask for 2 appointments, 1 day of hospitalization and a check-up. I have 2 jobs to get by financially. I have experienced an abortion this way before, it was traumatic. I’d rather be at home and manage my own abortion.

We found that compared to women over 36 years old, younger women who are 18-25 years old are two times more likely to find at-home abortion via telemedicine empowering and three times more likely to prefer having someone with them during the procedure compared to women 36 years old and above. They are, however, also two times more likely to perceive abortion stigma and 53.5% more likely to need to keep their abortion secret from their family or partner. Finally, younger people are also two times more likely to encounter financial difficulties while accessing abortion care. A young woman’s email to WoW demonstrates some of these difficulties:

> I called to make an appointment with the public hospital, and I was notified that the cost for an abortion is 630 Euros, of which I am totally not within the financial means to handle. I am a foreign student in France and because of the complications due to COVID, I’m currently in the process of renewing my student visa. However, my current visa is expired. Because of this as well, I don’t have access to the French national healthcare and unfortunately I am not insured in France. My parents who are my main financial support are very conservative and would not support paying that much for the abortion.

Even when abortion care is available and supposedly accessible, women reported personal circumstances, which prevents them from accessing care. These circumstances entailed being immigrant, wanting to keep abortion secret, experiencing financial constraints, abuse, or living in a controlling environment. Childcare commitments, lack of time due to work or school, and not having health insurance are also mentioned among obstacles women encounter in accessing local care. A woman in such a situation wrote: “I know abortion is legal in France, but I will tell you why I will not be able to have an abortion here. My companion is a violent man, I will never be able to have the opportunity to go to a hospital or a centre, without him watching me.” Another woman wrote in a similar vein:

> I live with an extremely conservative family and currently, we are quarantining due to coronavirus. I have no means of leaving my house and anytime I do leave my house, my whereabouts are strictly monitored. If I were to tell anyone in my family I was pregnant, my literal safety could be in jeopardy. I do not know what to do. I’m scared.

Women also mentioned encountering stigma and judgement. Some women had past traumatic experiences and hence indicated their preferences for telemedicine abortion at home. One woman wrote: “I have experienced an abortion in clinic before and it was traumatic. I’d rather be at home and go later for a check-up.” Another mentioned fear of judgement and pressure from medical staff:

> Working in a hospital myself and having had abortion 2 years ago, I am really afraid of being judged. In my region, getting an abortion is an obstacle course. The city gynaecologist does not perform abortions and doctors here rather pressure you to continue with pregnancy.

## Strengths and Limitations of the Study

The WoW dataset consists of self-reported data. The research made use of all available data WoW had from France in 2020; the analysed sample is not representative and the data was not randomized. The data for France was collected only for 2020, and therefore it does not allow a longitudinal multi-year analysis. The content analysis could have been enriched with in-depth qualitative interviews.

Despite these limitations, the study offers a significant contribution to the literature on abortion access in France during the pandemic. To our knowledge, there is no study on the demand for and drivers of telemedicine abortion in France. Given the ongoing health emergency, the study has a significant potential to inform policy makers on telemedicine abortion care provision within the French context but also in other similar contexts. Previous research conducted with the same dataset for other countries also provides us with the opportunity to compare the French case with other countries [19].

## Discussion

The COVID-19 outbreak has stirred up a health emergency worldwide. Related lockdown measures and travel restrictions have particularly affected access to abortion care, whose provision already largely occurs under conditions of structural violence due both macro-level constraints and the everyday micro challenges [20]. In fact, rather than creating new challenges, COVID-19 pandemic has illuminated and further amplified precipitating malfunctions in abortion care provision [20]. According to the Department of Research, Studies, Evaluation and Statistics (DRESS), 232.200 abortions were performed in France in 2019 [21]. Medical abortions constituted the 69% of all abortions and 25% among them were performed outside of hospital settings, in private offices of doctors and midwives and/or family planning centres [21]. DRESS notes, however, that recourse rates vary depending on the region; from 11.8 per 1,000 women in Pays de la Loire in metropolitan France to 39 per 1,000 women in overseas territories like Guadeloupe and Guyana [22]. It is estimated that each year 3 000 to 5 000 women travel abroad to have an abortion, notably to Spain and to the Netherlands, because they pass the permitted gestational limit in France [22].

Recourse to and drivers of induced abortion, as well as women’s experiences with local abortion care or travelling abroad to seek care, have been extensively studied within the French context [23, 24]. Expanding on this literature, this article provides initial insights into the drivers of telemedicine abortion in France by analysing women’s motivations and perceived barriers of access to abortion care. Our study shows that while the COVID-19 and related pandemic restrictions constituted an important push factor for women to choose telemedicine, the drivers of telemedicine are multidimensional and go beyond conditions unique to the pandemic. Our research findings suggest that telemedicine can help meet women’s needs and preferences for secrecy, privacy and comfort, while facilitating increased access to and enabling a more person-centred abortion care. Given the inequality of access, along with challenges women face every day, telemedicine can also help extend access to abortion in places where it remains limited, including rural areas. Improving access and adopting a person-centred approach to abortion care are likely to benefit those who are most vulnerable, living under resource-poor or financially dependent circumstances, grappling with higher rates of stigma and judgement [25].

In the context of the pandemic, several studies attest to increased demand for abortion via teleconsultation [26] [27]. Our research also contributes to these studies by demonstrating trends from France during the pandemic in general and lockdowns in particular. Moreover, our research findings provide a ground for comparison on the drivers of telemedicine in France with that of different countries. We found it important to note, for example, that compared to previous research conducted with WoW data on Germany in 2019, cost and stigma are much less of a driver in France (respectively 20.2% and 11%) than in Germany (respectively 37.4% and 40.2%) [28].

Examining some immediate impact of telemedicine abortion provision in the United Kingdom, a recent study found that introduction of telemedicine for abortion care in the has reduced the waiting times for termination by 4.2 days and more abortions were provided at less than 6 weeks gestation with telemedicine [29]. Further research should examine the impact of telemedicine abortion provision on the French health system to better inform COVID-19 response and policymaking during and beyond the pandemic.

## Conclusion

The demand for at-home medical abortion via teleconsultation increased in France during the lockdowns. However, drivers of telemedicine abortion are multi-dimensional and go beyond the conditions unique to the pandemic. Given the various constraints women continue to encounter in accessing safe abortion, telemedicine can help meet women’s preferences and needs for secrecy, privacy and comfort, while facilitating improved access to and enabling more person-centred abortion care.

## Data Availability

Data are available upon reasonable request.

## Notes

### Competing Interest Statement

Coauthors RG and HA work for or are affiliated with Women on Web.

### Author Declarations

The study was approved by the Regional Ethics Committee, Karolinska Institutet, Dnr 2009/2072-31/2 and Dnr 2020/05406.

